# Radioprotective effects of parthenolide lotion against skin injury caused by radiation (REPAIR-1): protocol for a phase 0, non-randomised, double-blind, interventional pilot study

**DOI:** 10.1101/2025.05.27.25328458

**Authors:** Connor M D Williams, Katherine L Morel, Agnieszka Kumorkiewicz-Jamro, Daniel P Johnstone, Hien V Le, Christopher J Sweeney, Hannah R Wardill

## Abstract

**Introduction:** Radiation forms the basis of many successful anti-cancer treatment regimens, killing tumour cells by irreversibly damaging their DNA. Unfortunately, radiation indiscriminately kills healthy cells, causing a range of debilitating and highly impactful side effects. One of the most common side effects is damage and burning of the skin, termed radiation-induced dermatitis (RID), which occurs in up to 95% of people undergoing radiation therapy. Although common, RID cannot be prevented and is inadequately managed with various topical solutions. Parthenolide has emerged as a promising radioprotective agent for the management of RID. The REPAIR-1 study is a phase 0, prospective, non-randomised, double-blind, interventional pilot study which aims to assess the efficacy of topical parthenolide application in limiting severe RID.

**Methods and analysis:** The REPAIR-1 study will be conducted at the Royal Adelaide Hospital, South Australia, Australia. Eligible participants for the REPAIR-1 study include adults diagnosed with locally advanced cancer of the head/neck scheduled to receive definitive, bilateral radiation therapy. Once enrolled, eligible participants will receive application of a 1% parthenolide, or placebo lotion, to opposite sides of the head/neck region before and after daily radiation therapy for the duration of their treatment course (6-7 weeks). The primary outcome of this study is absolute RID severity after 4 weeks of radiation therapy defined by both CTCAE v5.0 criteria and the RISRAS tool.

**Ethics and dissemination:** This protocol has been approved by Central Adelaide Local Health Network Human Research Ethics Committee (2024/HRE00241). All participants will be required to provide written informed consent. Results will be disseminated in peer-reviewed journals, and at scientific conferences.

**Trial registration number:** ACTRN12625000456459

## Introduction

Radiation therapy remains a mainstay in a wide range of cancer treatment protocols including, but not limited to, breast, prostate and head and neck cancer (HNC). Given the emergence of supportive cancer care as key priority for patients living with or beyond a cancer diagnosis, finding new ways to prevent the plethora of side effects experienced throughout standardised radiation regimens is of urgent need. Radiation induced dermatitis (RID) is a common sequela of radiation therapy, with upwards of 95% of patients experiencing RID to some extent. RID associated cracking of the skin is a major risk factor for infection and is extremely painful, causing immense distress for patients, their family and healthcare team. RID manifestation results from the indiscriminate, and localised damage to healthy cells, involving the generation of reactive oxygen species (ROS), a decrease in functional stem cells, initiation of epidermal and dermal inflammatory responses, and skin cell necrosis [1]. Currently, there are no universally approved ways to prevent or treat RID, and the only way to avoid it from worsening is to reduce the dose or frequency of radiation, thus decreasing the chance of cancer control. Hence, there is a pressing need for innovative and effective solutions to protect the skin during radiation therapy.

Given its prevalence, numerous efforts have evaluated the use of radioprotective compounds for the prevention of radiation mediated damage of healthy tissue, which have largely been limited by their toxicity and/or inability to differentiate between healthy tissue and malignant cells [2-6]. Parthenolide, a major active compound which is derived from the feverfew plant (*Tanacetum parthenium*), has emerged as a promising candidate, with its analogue dimethylaminoparthenolide (DMAPT) exhibiting preclinical radioprotective efficacy [7]. Notably, parthenolide has also been described to increase radiosensitivity in malignant cells [7, 8]. This differential radioprotection is attributed to redox-mediated modification of the Kelch-like ECH-associated protein 1/ Nuclear factor erythroid 2-related factor 2 (KEAP1/Nrf2) pathway, a prominent cellular defence mechanism against oxidative stress [9]. Thus, parthenolide activates nicotinamide adenine dinucleotide phosphate (NADPH) oxidase in malignant cells, driving further oxidative stress, whilst preserving redox homeostasis in healthy tissue through KEAP1/Nrf2 mediated adaptive oxidative responses [10].

Here, we present a protocol which aims to investigate the efficacy of parthenolide, when utilised as a topical lotion, for the prevention and management of RID. This protocol focuses on HNC patients scheduled to receive definitive, bilateral radiation to the head/neck region. This unique cohort provides opportunity for participants to act as their own internal control, given the bilateral nature of their treatment. We anticipate topical parthenolide application will reduce RID severity, and will be well tolerated by patients, positioning parthenolide as a strong therapeutic candidate for inclusion in the standard of care provided to people with HNC.

### Hypotheses

**Hypothesis 1**: Parthenolide will be well tolerated by people undergoing radiation for HNC

**Hypothesis 2**: Parthenolide will reduce the severity of RID in people with HNC

**Hypothesis 3**: Parthenolide will reduce the incidence of severe RID (CTCAE v5.0 Grade 3+) in people undergoing HNC radiation therapy

## Methods and analysis

### Study objectives

**Table 1.** Study outcome measures and assessment time points.

Study objectives:
| Table 1: Study outcome measures and assessment time points |  |  |
| --- | --- | --- |
| Objectives | Outcome measure(s) and method of collection | Assessment frequency/timepoints |
| Determine the incidence and severity of RID in study cohort, and compare intra-individual trajectories | RID assessed by CTCAE v5.0 criteria and the Radiation-induced Skin Reaction Assessment Scale (RISRAS) | Weekly and as needed per nurse and radiation therapist during treatment |
| Determine participant adherence to the investigational product | Number of investigational product (IP) treatment days missed in 6-7-week radiotherapy treatment | Daily during IP administration period (recorded by nurse, radiation therapist or trial coordinator) |
| Determine rate of treatment-emergent adverse events (TE-AEs) in study cohort | Rate of Adverse events (AEs) probably or definitely linked to the IP assessed by member of the study team using CTCAE v5.0 criteria | Weekly during IP administration or *ad hoc* as they present |
| Determine use of standard of care skin care | Participant self-reported use frequency | Weekly during IP administration period |
| Determine participant preference of the investigational product vs control product | Proportion of participants that prefer IP to comparator determined by binary question (i.e. "which intervention did you prefer, left or right side?") | At completion of radiotherapy and IP |

### Trial design

The REPAIR-1 study is a Phase 0, prospective, non-randomised, double-blind, interventional pilot study (N=10 participants) with internal controls. The study will exclusively operate and recruit patients from the Royal Adelaide Hospital (RAH), South Australia (SA), Australia.

### Intervention and placebo

Eligible participants (Table 2) will act as their own internal control (Figure 1), receiving both IP and placebo lotions. One side of the head/neck will receive a proprietary base solution. Parthenolide has been extracted using an ethanol extraction process at University of Adelaide (UoA) from *Tanacetum Parthenium (feverfew) grown at Roseworthy campus, (UoA)*. Preliminary *In vitro* dosing of radiation has shown radiation protection of normal cells is achieved with 3 to 10 μM of parthenolide which equates to approximately 1% solution in this setting. Therefore, a concentration of 1% parthenolide will be used throughout the study. The opposite side of the head/neck will receive the base solution with no additional components.

**Table 2.**
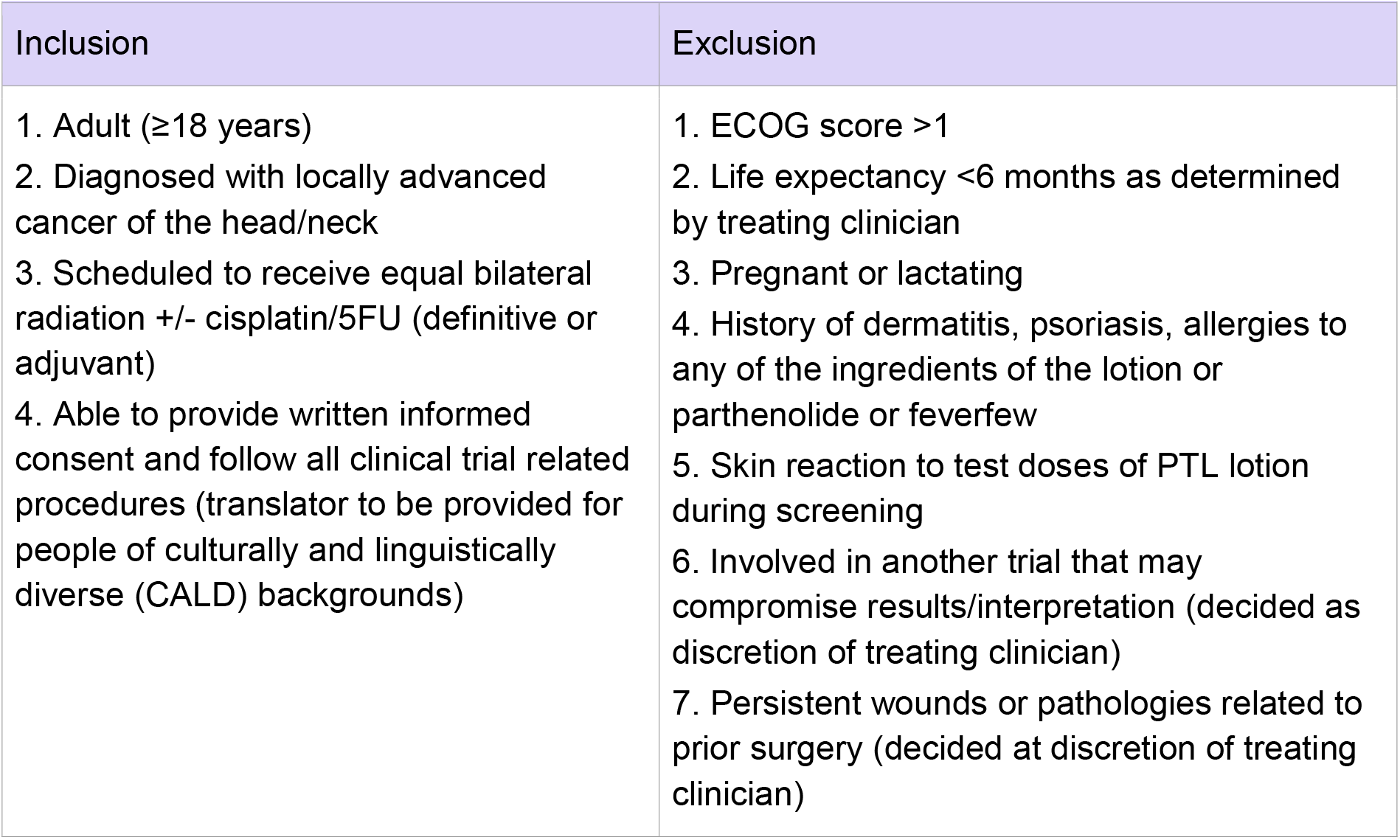
Inclusion and exclusion criteria.

**Figure 1.**
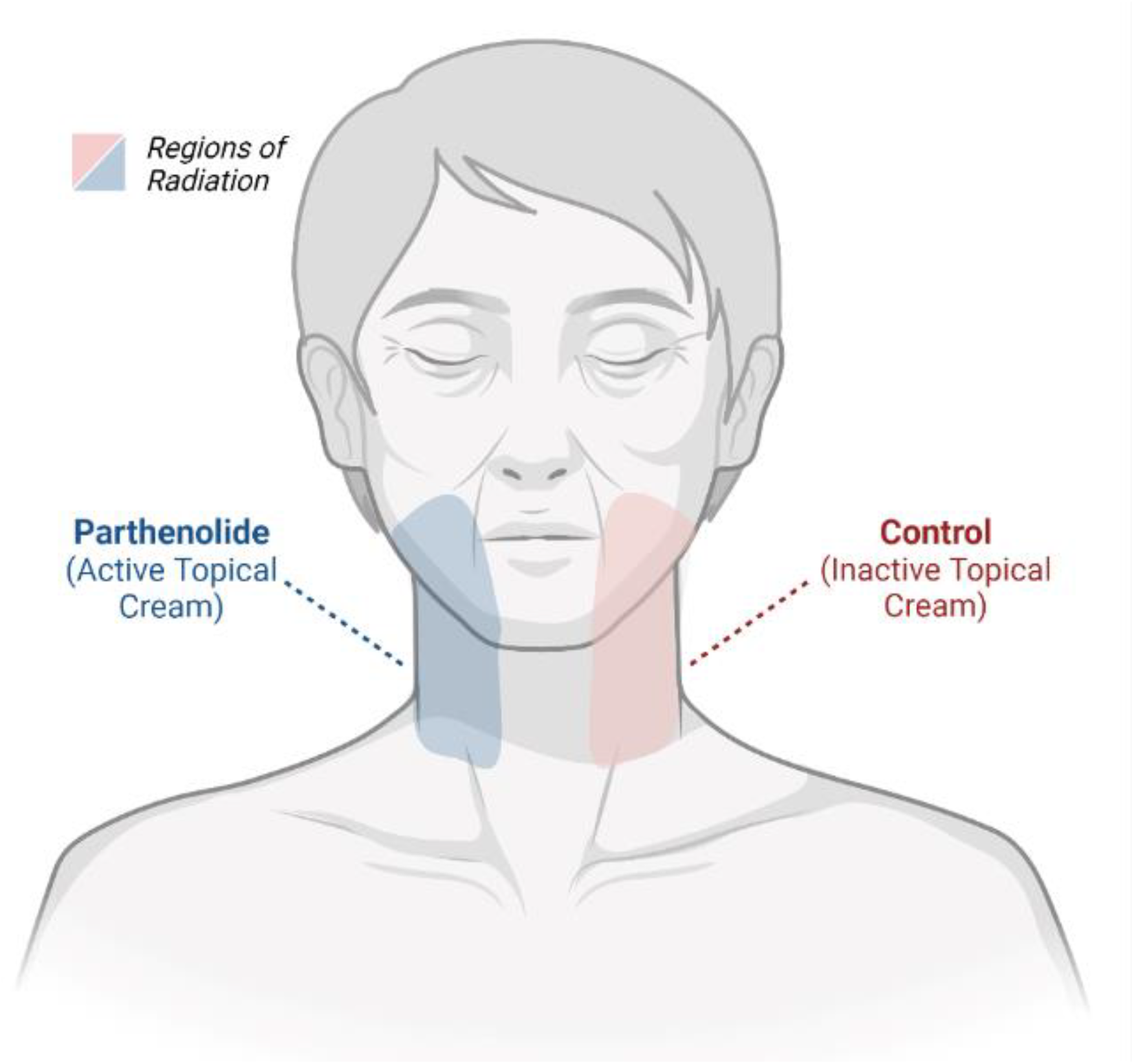
Experimental design with participants using both the active IP and comparator on different sides of the face/neck Created with BioRender.com

### Randomisation and double-blind conditions

This is a double-blind study, with the products only identified as LEFT and RIGHT. Packaging will be identical between IP and placebo lotions, manufactured by Plantworx. The unblinded monitor, who will not be involved in the study assessments or application of the lotion, will be the only person to know the allocation of the IP to LEFT or RIGHT. The unblinded monitor will have no interactions with study participants and will not be involved in any study assessments.

### Recruitment and consent

Potentially eligible participants will be identified at RAH multidisciplinary team meetings by a member of the RAH radiation oncology unit trials team and the study will be introduced to the participant by their clinician at the earliest possible convenience. If willing, interested and eligible, a follow up appointment will be scheduled for the participant. At this appointment, eligibility will be established and written consent will be provided. Once consented, the participant will be given a unique ID number which will be used to identify all study material.

### Timing and dose

All participants will be provided with the IP and placebo lotions prior to starting radiotherapy (at time of mask fitting), and instructed to test the cream on a small patch of skin (on the arm) for 3 days with photos taken for review. Left arm will receive the LEFT labelled lotion and the right arm will receive the RIGHT labelled lotion. If any reactions are reported or observed, the participant will be withdrawn. Eligible participants that do not develop any reactions during the patch test will have both products applied (figure 1), before and after each radiotherapy, with application continuing until completion of their radiotherapy protocol. The duration is 6-7 weeks, dictated by the duration of their radiation protocol.

Lotions will be stored on site at the RAH at 4°C. A nurse will instruct the participant in applying the LEFT and RIGHT lotions to each side of the face. They will observe and document lotion application. Participants will be permitted to use standard of care (e.g. sorbolene) ad libitum, and asked to report on this at their weekly assessments.

### Study Assessments

After consenting to the study, participants will undergo a baseline assessment which will record date of birth, sex, height, weight, diagnosis, treatment specifications, comorbidities, concurrent medications and ECOG performance status. People undergoing radiation therapy are required to visit hospital Monday-Friday for daily radiation for 6-7 weeks. As such, all assessments will be performed in alignment with these frequent visits.

The following assessments will be performed weekly for the duration of participants treatment schedule:

- CTCAE assessment for AEs
- CTCAE assessment for RID
- RISRAS assessment for RID
- Adherence evaluation
  ∘ Staff applying the lotion will be asked to report the number of days within the past week in which the IP or control product was not applied, and the reason(s) why.
  ∘ An ongoing record will be kept by staff at the Royal Adelaide Hospital to aid in recording daily use of the IP as well as an opportunity to record any adverse reactions indicated by the participant.
- Photos of region receiving radiation and lotion application

### Primary outcomes

- Absolute RID severity at week 4 defined by CTCAE v5.0 criteria
- Absolute RID severity at week 4 defined by RISRAS tool

### Secondary outcomes

- Incidence of <u>severe</u> RID (defined as CTCAE v5.0 grade 3+) at week 4 of treatment
- Incidence of <u>severe</u> RID (defined as CTCAE v5.0 grade 3+) at each week of treatment (week 0-7)
- Average RID grade defined by CTCAE v5.0 each week from week 4-7
- Average RID score defined by RISRAS each week from week 4-7
- AUC of RID grade (defined by CTCAE v5.0) for full treatment period (week 0-7)
- AUC of RID grade (defined by RISRAS) for full treatment period (week 0-7)
- Time until RID (CTCAE v5.0 grade 1+) development
- Rate of TE-AEs
- Adherence to IP
- Participant preference to IP vs comparator
- Cumulative dose of radiation to each side of the head/neck
- Use of standard of care product(s) (Sorbolene/QV cream) on each side of the face, as reported by the participant

### Biospecimen collection

Skin samples will be collected non-invasively from participants using D-Squame Standard Sampling Discs. A maximum of 20 discs, repeatedly applied and removed on the same region of skin will be pressed with a pressure of 225 gcm^2^ for 10 seconds at a single timepoint (on each side). Skin will be collected at baseline and after 3 weeks of radiation. This time point has been selected as it is before RID becomes severe, hence avoiding skin collection when the skin barrier is compromised.

### Discontinuation/modification and adherence

People with HNC are at risk of drop out due to the intensity of the radiation treatment they receive. The burden of side effects and the requirements of treatment (i.e. daily visits to the hospital) are high, however, this also makes them a receptive audience for trials that aim to reduce side effect burden. The frequency at which they are already required to attend the hospital for their radiation also reduces the burden of trial participation, as such, a dropout rate of 10% is expected. The trial will continue to recruit until N=10 participants have successfully reached the primary endpoint of 4 weeks. Any drop outs prior to this will be captured and recorded as trial data (assuming the participant consents to their data being used after their withdrawal), however, if they did not reach the end of treatment (week 4 of radiation) for any reason other than skin toxicity prior to week 4, they will not be included in any analyses related to the primary objective of the trial. However, their data from the weekly assessments will be tabulated and rate and degree of RID for control versus treatment as well as intra-patient evaluations will be performed.

Week 4 has been chosen as the primary endpoint as most patients receive a comparable dose to each side of the neck at this time-point, with less chance of dose modifications which could modify analysis. Outcomes beyond week 4 are to be reported as detailed above and potential confounders such as radiation dose modification noted.

### Adverse events

People undergoing radiotherapy will develop a range of AEs related to their cancer treatment not the IP. Hence, all AEs will be reviewed within 72 hours, and the site PI will determine their relationship to the IP. If deemed probably or definitely related to the IP, these AEs will be defined as a treatment emergent AE (TE-AE). AEs will be identified during weekly assessments using criteria established by the NCI CTCAE, V.5.0. Where parameters are not addressed within the criteria, severity of AEs should be graded as:

- Mild: Aware of sign or symptom, but easily tolerated
- Moderate: Discomfort enough to cause interference with usual activities
- Severe: Incapacitating with inability to work or perform usual activities
- Life-threatening: Participant is at immediate risk of death
- Fatal: Death

Suspected unexpected serious adverse reactions (SUSARs) are adverse reactions that are untoward and unintended responses to the IP (i.e. a serious but unexpected TE-AE).

Unexpected adverse reactions that are classed as suspected unexpected serious adverse reactions (i.e. a serious but unexpected TE-AE). Unexpected adverse reactions are SUSARs if the following three conditions are met:

- The event must be serious
- There must be a certain degree of probability that the event is a harmful and an undesirable reaction to the medicinal product under investigation, regardless of the administered dose
- The adverse reaction must be <u>unexpected</u>, that is to say, the nature and severity of the adverse reaction are not in agreement with the product information as recorded in the:
  ∘ Summary of Product Characteristics (SPC) for an authorized medicinal product
  ∘ Investigator’s Brochure for an unauthorised medicinal product
  ∘ All SUSARs will be reviewed by the Safety Monitoring Committee and documented in the CRF. The Safety Monitoring Committee will determine the appropriate course of events.

If the Safety Monitoring Committee identifies a SUSAR, they have the discretion to stop the trial or withhold the IP from the participant. This will be discussed and determined by the Safety Monitoring Committee depending on the severity of the AE and its relationship with the IP.

### Study assessment schedule

**Table 3.**
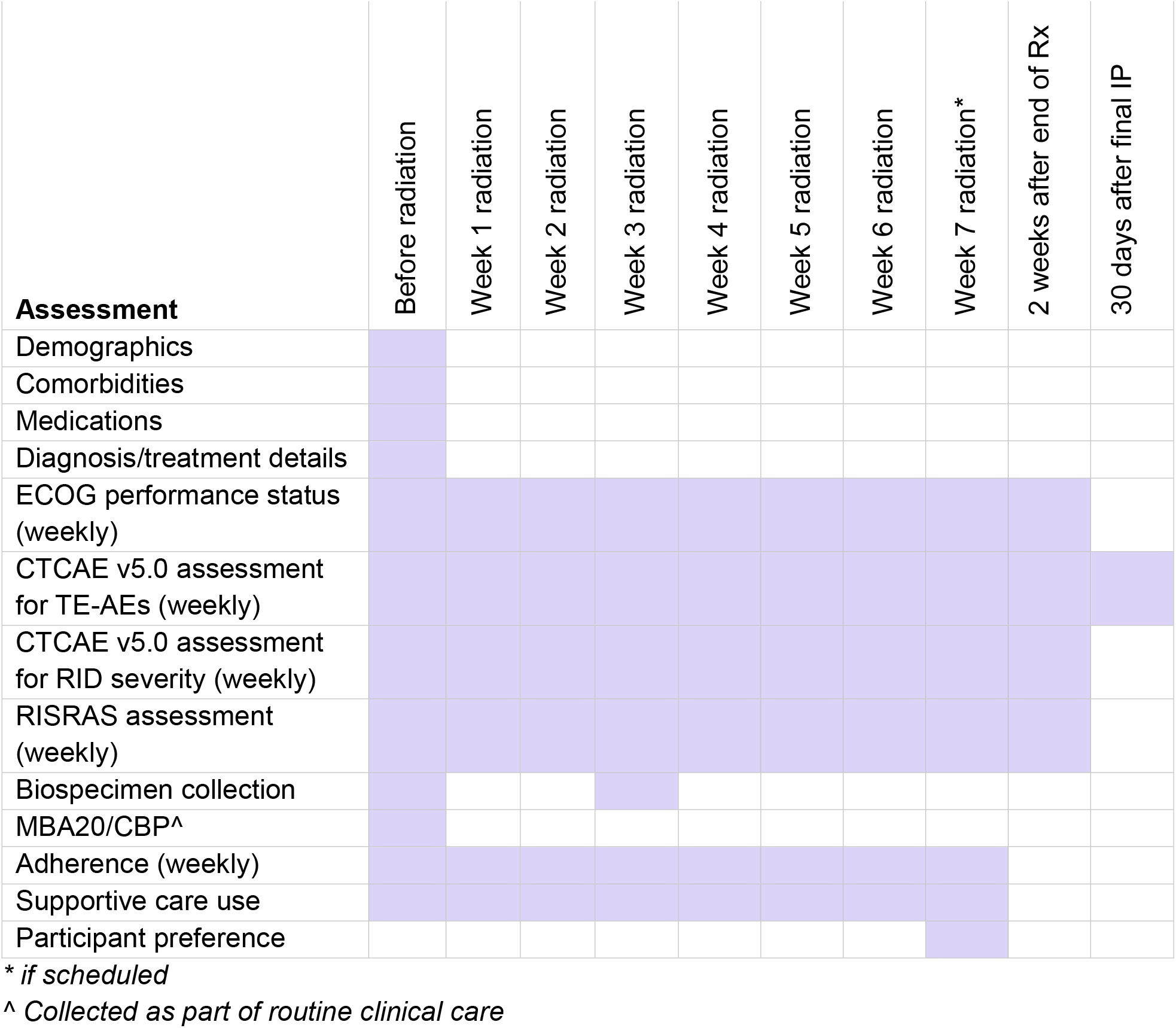
Study assessment schedule.

### Trial oversight, monitoring, auditing and data management

The REPAIR-1 study is a collaboration between the University of Adelaide, the South Australian immunoGENomics Cancer Institute (SAiGENCI), Plantworx and the Central Adelaide Local Health Network (study sponsor). All data collected for each participant will be kept in a trial file, which will contain the CRFs. Any corrected and amended data, copies of AE reports, file notes, pathology reports, medication chart for all trial participants. All material will be de-identified. Consent forms will be stored separately. Any participant hard copy files will be securely stored in a locked filing cabinet that is only accessible to study staff and away from the administrative files for the study. Electronic files are stored on password protected files in secure offices and all study files will be stored for 15 years.

Identifiable data (e.g. pathology reports) will be de-identified and filed with the study documents. All participant files will be reconciled and stored along with all study materials – both hard copy and electronic – consistent with ICH GCP and applicable regulations regarding the retention and disposal of participant records.

All hard copy study data (de-identified), e.g. CRFs, will be scanned into electronic PDFs and stored on a secure, web-based platform (LabArchives) before being entered into REDCap; a secure web-based application designed to support data capture for research studies, providing user- friendly web-based case report forms, real-time data entry validation, audit trails and non-identified data export mechanism to common statistical packages. All web-based information transmissions in REDCap are protected via Secure Sockets Layer (SSL) encryption (data entry, survey submission, web browsing, etc).

### Data analysis and dissemination

All qualitative data will be presented as descriptive data (e.g. participant demographics) and compared between groups using a Chi squared test. Study endpoints will be compared using paired t-tests and/or Chi squared tests. The interpretation and analysis of results will be performed by the investigators and their delegates. Results will be published in appropriate peer-reviewed journals and presented at national and international scientific meetings (e.g. Trans-Tasman Radiation Oncology Group, Multinational Association for Supportive Care in Cancer, Clinical Oncology Society of Australia). We will also work with community organisations such as Cancer Voices SA to ensure these data are communicated with consumers.

### Ethics

The study protocol was approved by the Central Adelaide Local Health Network Human Research Ethics Committee (2024/HRE00241). This Protocol has been designed to comply with the Declaration of Helsinki and any subsequent amendments, the ICH Guidelines for Good Clinical Practice (CPMP/ICH/153/95) annotated with TGA comments (July 2000), the NHMRC National Statement on Ethical Conduct in Research involving Humans 2023, the policies and procedures of any applicable local guidelines. The trial will be conducted in compliance with the Protocol, International Council for Harmonisation, Good Clinical Practice (ICH GCP) Guidelines in Australia, and applicable regulatory requirements. This trial has been registered with the Australian and New Zealand Clinical Trials Registry and Therapeutic Goods Administration.

When potential participants are introduced to the trial, they will be informed of the study objectives and that participation is voluntary. Each participant will be required to provide written consent.

## Discussion

With radiation heavily embedded in standardised anti-cancer protocols for a range of localised cancers, there remains urgent need for improved management of the associated and debilitating off target toxicities. Despite previous efforts to protect the skin from radiation damage, there remains no universally approved strategies to so. The REPAIR-1 study will establish the feasibility and efficacy of topical parthenolide application as a radioprotective agent, aiming to offer a mechanistically novel approach to prevent RID whilst still ensuring radiation efficacy.

The REPAIR-1 study targets patients who are scheduled to receive bi-lateral radiation to the head or neck region. This unique cohort of patients provides opportunity for internal controls, with participants receiving the IP and vehicle base lotion simultaneously. In addition to the use of clinically validated tools for the grading of RID severity, this will allow patients to record personal preference at the cessation of treatment. With growing demand from patients for enhanced supportive cancer care strategies, such outcome measures will prove vital in facilitating future clinical guidelines for the management of RID.

## Data Availability

All data produced throughout the present study are available upon reasonable request to the authors

